# A non-*APOE* Polygenic score for Alzheimer’s disease and *APOE-ε4* have independent associations with dementia in the Health and Retirement Study

**DOI:** 10.1101/2020.02.10.20021667

**Authors:** Kelly M. Bakulski, Harita S. Vadari, Jessica D. Faul, Steven G. Heeringa, Sharon LR Kardia, Kenneth M Langa, Jennifer A. Smith, Jennifer J. Manly, Colter M. Mitchell, Kelly S. Benke, Erin B. Ware

**Author notes:** **Corresponding Author:** Erin B. Ware, University of Michigan, Survey Research Center, Population, Neurodevelopment and Genetics, 426 Thompson St, Ann Arbor, MI, 48104, USA.

## Abstract

**INTRODUCTION:** Alzheimer’s disease (AD) is a common and costly neurodegenerative disorder. A large proportion of risk is heritable and many genetic risk factors for AD have been identified. The cumulative genetic risk of known markers has not been benchmarked for dementia in a population-based sample.

**METHODS:** In the United States population-based Health and Retirement Study (HRS) (waves 1995-2014), we evaluated the role of cumulative genetic risk for AD, with and without the *APOE-ε4* alleles, on dementia status (dementia, cognitive impairment without dementia, borderline cognitive impairment without dementia, cognitively normal). We used logistic regression, accounting for demographic covariates and genetic principal components, and analyses were stratified by European and African genetic ancestry.

**RESULTS:** In the European ancestry sample (n=8399), both AD polygenic score excluding the *APOE* genetic region (odds ratio (OR)=1.10; 95% confidence interval (CI): 1.00, 1.20) and the presence of any *APOE*-*ε4* alleles (OR=2.42; 95% CI: 1.99, 2.95) were associated with the odds of dementia relative to normal cognition in a mutually-adjusted model. In the African ancestry sample (n=1605), the presence of any *APOE*-*ε4* alleles was associated with 1.77 (95% CI: 1.20, 2.61) times higher odds of dementia, while the AD polygenic score excluding the *APOE* genetic region was not significantly associated with the odds of dementia relative to normal cognition 1.06 (95% CI: 0.97, 1.30).

**DISCUSSION:** Cumulative genetic risk for AD and *APOE-ε4* are both independent predictors of dementia. This study provides important insight into the polygenic nature of dementia and demonstrates the utility of polygenic scores in dementia research.

## Background

Dementia is a neurodegenerative disorder characterized by progressive cognitive decline, which leads to a loss of independence and causes medical, social, and economic burdens on the population. With the population of those 65 and older estimated to grow from 55 million to 88 million between 2019 and 2050, the number of people with dementia is likely to increase (1, 2). In 2019, the estimated health care, long term care, and hospice care costs associated with dementia is $290 billion (1, 3). Identifying risk factors for dementia is essential to prevention and potential treatment. Heritability estimates for dementia attribute 50-80% of risk to genetic factors (4). Additionally, many environmental factors have been identified with varying degrees of risk on dementia (5). With no present cure for dementia, understanding etiologic and preventive measures is essential to reducing the burden of disease on individuals as well as the population.

The most common dementia genetic susceptibility locus is in the Apolipoprotein E (*APOE*) gene, represented by multiple alleles: *APOE ε2, APOE ε3, APOE ε4*. These combined *APOE* alleles (ε2/ ε2, ε2/ ε3, ε2/ ε4, ε3/ ε3, ε3/ ε4, or ε4/ ε4) confer either a protective effect against AD (e.g. ε2/ ε2) or an increased risk of AD (e.g. ε4/ ε4) (6). In the Rotterdam study, those with the *APOE* ε4/ ε4 genotype had 11.2 times higher odds of dementia (95% Confidence Interval (CI: 3.6-35.2), compared to the ε3/ ε3 genotype (7). Those with the *APOE* ε3/ ε4 genotype had 1.7 times higher odds of dementia (95% CI: 1.0-2.9), compared to ε3/ ε3 (7). In an African-American study sample, those with the *APOE* ε4/ ε4 genotype had 10.5 higher odds of dementia (95% CI: 5.1-21.8), relative to those with ε3/ ε3 (8). In the same study, those with only a single copy of the ε4 allele had 2.6 higher odds of dementia (95% CI: 1.8-3.7), relative to those with ε3/ ε3 (8). The presence of *APOE* ε4 alleles increase risk for dementia, though they are neither necessary nor sufficient for disease and they do not fully capture the complex, polygenic nature of dementia.

Among the main dementia sub-types (Alzheimer’s, vascular, frontotemporal, Lewy body, and mixed) (9), Alzheimer’s disease (AD) is the most common (70% of cases) (10). Strong, Mendelian effects are observed for rare genetic loci in early-onset AD (mutations in the amyloid precursor protein (*APP*), presenilin 1 (*PSEN1*), and presenilin 2 (*PSEN2*) genes) (11). Late-onset AD is the more common and sporadic form of AD. In addition to the *APOE* locus, many other genetic sites are associated with late-onset AD, identified through genome-wide association studies (GWAS) (12). Cumulative genetic risk for complex outcomes, like AD, can be summarized using polygenic risk scores (PGS), based on *a priori* knowledge of the genetics for that trait (13). PGS are constructed by weighting single nucleotide polymorphisms (SNPs) by their known association with the trait from the GWAS, and summing them into a single, per-person, cumulative PGS measure, which is assumed to be additive. In a recent non-Hispanic White, familial AD study sample, an unweighted PGS was constructed using the number of participant risk alleles at 19 genome-wide significant AD SNPs, and a one-standard deviation unit increase in the PGS was associated with 1.29 times increased odds of clinically diagnosed late-onset AD (95% CI: 1.21-1.37), relative to unaffected family members (14). While PGSs using genome-wide significant SNPs may be more biologically interpretable, the use of all SNPs (i.e. a genome-wide score) often explains significantly more of the variation in an outcome than a score using only genome-wide significant SNPs (15, 16).

We investigated whether cumulative genetic risk for AD – over and above the risk already established by the *APOE* ε4 allele – is associated with odds of dementia. We newly estimated cumulative genetic risk for AD using an AD PGS that incorporates SNPs across the entire genome. In a unique, large, population-based study, the Health and Retirement Study (HRS), we characterized the utility of PGS in a European genetic ancestry sample. We additionally evaluate an AD PGS in an African genetic ancestry sample, where the PGS may have value, albeit as a less informative instrument given the European-based PGS weights. By evaluating the role of cumulative genetic risk on dementia status, we provide important insight into the genetic correlates of dementia and demonstrate the utility of AD PGSs in dementia research across ancestries.

## Methods

### Health and Retirement Study (HRS) sample and design

The HRS is a nationally representative, longitudinal panel cohort study of adults over the age of 50 (n∼22000, per wave) in the United States. Detailed methods are described elsewhere (17). The HRS was collected biennially beginning in 1992, through face-to-face interviews, mail-in surveys, and leave-behind questionnaires, with dedicated collection efforts to obtain saliva (in 2006 and 2008), dried blood spots (2006-2012), genotyping (2006-2012), and venous blood draws (in 2016) for biomarker data and genetics. This analysis included participants from ten waves (1995-2014). The HRS is sponsored by the National Institute on Aging (U01AG009740) and is conducted by the University of Michigan. Informed consent was obtained from all participants and the University of Michigan Institutional Review Board approved these analyses (HUM00056464).

### Health outcomes: wave specific dementia status and summary cognition status

Langa-Weir cognition status at each wave was defined using a method previously described and validated in the HRS (18, 19). A three-level cognition status variable (dementia, cognitive impairment-no dementia (CIND), normal cognition) was created from available survey instruments and imputed for those self-respondents missing cognitive tasks using multivariate, regression-based imputation and variance estimation (20). Categorization was performed separately for self-report and proxy respondents. This method was validated against a clinically evaluated subsample of the HRS where 76% of self-respondents and 84% of proxy respondents were correctly classified as having dementia (19).

Because cognition can fluctuate between waves (21) and we were interested in cumulative cognitive status, we constructed a summary measure of participants’ cognition status across all available visits. Each participant was assigned a summary cognition status based on their set of Langa-Weir cognition values across their entire time in the study (ranging from two to ten wave-specific measures) and placed into one of five possible summary statuses: dementia, CIND, borderline CIND, cognitively normal, and unclassified (**Supplementary Table 1**). Participants were excluded from summary cognition status classification if they had fewer than three eligible waves of cognition measured. Observations were excluded from summary cognitive classification if the participant was less than 60 years of age at the time of cognitive assessment.

Summary cognition status was based on the following criteria: first, participants with the same cognition status in their last two visits were given that cognition classification: dementia, CIND, or cognitively normal status (n=7756). Second, participants with a monotonic descent to dementia or fluctuations between CIND and normal cognition with the most recent visit categorized as dementia were classified in the dementia category (n=460). Third, participants with no dementia status in any wave, who fluctuated between CIND and cognitively normal, were classified as a new category, borderline CIND (n=1568). Next, participants with a majority (>50%) of normal cognition visits or CIND, with one dementia classification in the last two visits, were assigned CIND status (n=198). Next, participants with a majority of dementia classifications and one dementia status in their last two visits were classified as having dementia (n=22). Finally, participants with a normal cognition classification after a dementia classification or those who did not follow any of the above stated patterns were considered unclassified (n=171).

### Genetic risk for Alzheimer’s disease

Molecular data for the HRS was downloaded from dbGap (phs000428.v2.p2) which includes genetic data from samples obtained in 2006, 2008, and half of 2010. Genotype information was obtained from saliva DNA and genotyped on the Illumina Human Omni-2.5-4v1 and Illumina Human Omni-2.5-8v1 Quad BeadChip platform (22) at the Center for Inherited Disease Research. HapMap (23) controls were genotyped along with the study samples. Autosomal SNPs were filtered based on a missing call rate < 5% and minor allele frequency > 5%. SNPs were excluded if they were discordant between those in the HapMap controls and external HapMap data set (22). In addition, the highly variant 2q21 (lactase (*LCT*) gene), human leukocyte antigen (*HLA*) gene, 8p23, and 17q21.31 regions were excluded from the initial pool. Measured SNPs used for principal component (PC) analysis were selected by linkage disequilibrium pruning which selected 155,707 SNPs with all pairs having r^2^ < 0.1 in a sliding 10 Mb window from an initial pool consisting of all included SNPs.

The HRS genomic samples use a combination of both self-reported race/ethnicity and genetic ancestry to identify ancestrally homogenous analytic samples. Participants’ genetic ancestry was identified through PC analysis on independent genome-wide SNPs, in combination with self-reported race/ethnicity. PC analysis was conducted on unrelated study subjects and HapMap controls and each participant’s loading for eigenvectors (or PC’s) one and two were calculated. The European ancestry sample included all self-reported non-Hispanic White participants that were within ± one standard deviation of the mean for eigenvector one. The African ancestry sample included all self-reported non-Hispanic Black participants within two standard deviations of the mean of eigenvector one and ± one standard deviation of the mean for eigenvector two. Concordant genetic ancestry and self-reported race/ethnicity participants were retained (n=2279 non-Hispanic Black/African ancestry; n=9991 non-Hispanic White/European ancestry). Self-reported race/ethnicity and genetic ancestry are perfectly correlated by selection in this study, importantly eliminating our ability to test for effects in discordant or mixed racial/ancestral groups. To create sample eigenvectors for possible population stratification covariates, PC analysis was performed again within each ancestry sample. These values were released as a public data product by the HRS (22).

Two genetic variants (rs7412 and rs429358) contribute to the *APOE* isoforms. The measured genotype of rs7412 failed quality control thresholds and rs429358 was not available on the genotyping chip for part of the HRS sample using the Illumina Human Omni-2.5-4v1; therefore, the imputed values were used to identify *APOE* isoforms in the present study. One of six *APOE* isoforms (*ε2/ε2, ε2/ε3, ε2/ε4, ε3/ε3, ε3/ε4 and ε4/ε4)* was assigned to each individual using 1000 Genomes Project imputation data (worldwide reference panel of all 1,092 samples from the phase I integrated variant set (v3, released March 2012) (24). Our primary analysis included a binary indicator for all individuals based on the presence of the *ε4* allele in their genotype. Sensitivity analyses included additional categorizations for the *APOE* isoforms.

Cumulative genetic risk for AD was calculated using polygenic scores (22). AD PGSs were constructed using the following formula:

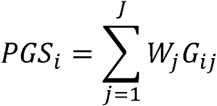

where *i* is individual *i* (i=1 to N), *j* is SNP *j* (j=1 to J), and *W* is the meta-analysis effect size for SNP *j. G* is the genotype, or the number of reference alleles (zero, one, or two), for individual *i* at SNP *j*. Imputed SNPs were not used in the AD PGS (22). Effect estimates were taken from the summary statistics from a large meta-analysis of Alzheimer’s disease GWAS in European ancestry (25). The stage 1 GWAS analysis, from which the weights were derived, included 63926 participants (n_cases_ =21982 and n_controls_ =41944). Summary statistics were obtained from National Institute on Aging Genetics of Alzheimer’s Disease Data Storage Site (https://www.niagads.org/datasets/ng00075). We sought to test the independent effects of AD PGS and *APOE* status; therefore, our AD PGS contains all SNPs that overlap between the genotyped data in the HRS and the AD GWAS summary statistics (25), after removing the region on chromosome 19 containing the linkage disequilibrium block of the *APOE* gene (chr19:45384477-45432606, GRCh37/hg19) from the summary statistics. Only those variants with P-values <0.01 in the summary statistics were included in the PGS, based on evaluations of PGS developed across multiple P-value thresholds and their degrees of association with the outcome (26).

Though the Kunkle et al., 2019 analysis was conducted in individuals of European ancestry, we conducted our analyses in both the European ancestry and the African ancestry cohorts. We note the HRS recommendation that “*PGSs for other ancestry groups may not have the same predictive capacity…”* (22, 27), and *“users (should) perform analyses separately by ancestral group and, at the very least, adjust for PCs 1-5*” (28). We emphasize the need for large GWAS on non-European ancestries with widely available summary statistics to help advance the knowledge in this field. AD PGS were z-score standardized within ancestry.

### Covariates

Information on sex (female, coded “0”) and number of years of education were collected at the start of HRS participation for each respondent. We considered the age at last cognitive assessment in our analyses as well as the year of the last assessment. In a sensitivity model, we include other risk factors for AD based on previously observed association. These factors include hypertension (29), history of diabetes (30), smoking behavior (31), alcohol use (32), BMI (33), depressive symptoms (34), and stroke (35). History of hypertension (no, coded “0”), diabetes (no, coded “0”), smoking (never, former, current), and alcohol use (never, coded “0”) were all assessed at the last cognitive visit using variables from the RAND Center for the Study of Aging, which is supported by the National Institute of Aging and Social Security Administration (36). If the last cognitive visit was face-to-face, we preferentially used the concurrent measured BMI (kg/m^2^) followed by the participant’s self-reported BMI at that wave. If these two values were missing, we selected measured BMI from the prior wave followed by self-reported BMI from the prior wave. Depressive symptoms, measured by the 8-item Center for Epidemiological Scales – Depression questionnaire, were averaged across all waves concurrent to and prior to the last cognitive measure (37). This value was then dichotomized at the ancestry-specific mean such that 1 represents higher than mean depressive symptoms and 0 is less than mean depressive symptoms. At each wave participants were queried on their history of stroke or transient ischemic attack. This information was used to construct a summary variable for ever having a stroke (no, coded “0”) at the last year of cognitive assessment.

### Statistical analysis

Analyses were performed using SAS 9.4 (SAS Institute, Cary, NC) and R (Version 3.5.1, R Foundation for Statistical Computing) (38). Code to complete analyses is available (https://github.com/bakulskilab). We calculated univariate descriptive statistics. Due to multiple covariates (sex, age, *APOE-ε4* status, education) violating the assumptions of proportional odds we used separate logistic regressions to model the odds of impaired cognition (dementia, CIND, or borderline CIND) with normal cognition as a reference category. All models were adjusted for age and year at last cognition measurement, sex, years of education, and two ancestry-specific genetic principal components. Analyses were stratified by ancestry (European and African). Stratification of these models is important given that genetic architecture varies by ancestry, the PGSs were created using weights derived from a European ancestry population (25), and risk factor profiles may not be the same across groups (39).

Our base model included age of last visit, sex, educational attainment, year of last visit, and two ancestry-specific genetic principal components (Model 1). Model 2 adds one of the AD genetic components (Model 2a: AD PGS, Model 2b: *APOE-ε4* status) to the base model. Both genetic components were included in Model 3.

### Sensitivity analyses

To assess the robustness of our findings to methodological and analytic decisions, we conducted several sensitivity analyses. First, to assess potential linear deviations among pairs of AD PGSs, we performed correlation tests (Pearson, ρ) comparing an AD PGS without variants in the *APOE* gene region (removing 444 variants from chr19:45384477-45432606, GRCh37/hg19 from the summary statistics; **Supplementary Table 2**), to one with variants in the *APOE* gene region. Second, to examine the effect of different P-value cutoffs for variants included in the AD PGS, we compared the performance of AD PGS developed using variable GWAS P-value cutoffs (pT=1, 0.3, 0.1, 0.05, 0.01, 0.001) in Model 2a. Third, to account for potential survival bias, similar logistic models were performed with the oldest HRS cohorts, Assets and Health Dynamics (AHEAD: birth year < 1924) and Children of the Depression Era (CODA: birth years 1923-1930) removed. Fourth, to test whether the effect of AD PGS was different in the presence of *APOE* and vice versa, we tested for a potential multiplicative interaction between *APOE* and the AD PGS. Fifth, to characterize the effect of the *APOE* locus, a set of logistic regression models examined a three-level *APOE* variable, based on the number of *ε4* copies in the individual (*ε2/ε2, ε2/ε3, ε3/ε3*, coded “0”; *ε2/ε4, ε3/ε4*, coded “1”; *ε4/ε4*, coded “2”). While we attempted to assess a third specification of the *APOE* locus where the protective *ε2/ ε2* haplotype and the deleterious *ε4/ ε4* were explicitly modeled compared to any other haplotype, we did not have enough individuals in the *ε2/ ε2* haplotype to proceed (<1% of the total sample in each ancestry). Sixth, to assess the potential effect of health behaviors on the relationships presented, we presented fully adjusted models, which included all variables from Model 3 as well as history of hypertension, diabetes, smoking, alcohol use, stroke, and depressive symptoms (Model 4).

### Receiver Operating Characteristic Curve

To evaluate the classification capabilities of the models, receiver operating characteristic (ROC) curves were estimated. Areas under the curve (AUC) for logistic models of dementia versus normal cognition for Models 1, 2a, 2b, 3, and 4 were compared. C-statistics from Models 2a, 2b, 3, and 4 were compared to Model 1 to assess whether the addition of AD PGS, *APOE- ε4* status, or the combination of AD PGS and *APOE-ε4* status improved the classification ability over that of the Model 1 using a chi-squared test from an ROCCONTRAST statement in the PROC LOGISTIC procedure. We further evaluated the AUC of Model 3 (with both AD PGS and *APOE-ε4* status) relative to Model 2b (Model 1 + *APOE-ε4* status) to determine if the addition of AD PGS significantly improved the classification of dementia and normal cognition over and above *APOE-ε4* status.

### Attributable fraction

To determine the proportion of the dementia burden that would be reduced in the absence of elevated cumulative genetic risk (highest twentieth percentile AD PGS or *APOE-ε4* allele carrier), we calculated the attributable fraction for the dementia vs normal cognition models. We first compared those in the highest twentieth percentile of AD PGS to those in the lowest twentieth percentile of AD PGS using an odds ratio from adjusted logistic regression models. Next, we compared those with at least one copy of the *APOE-ε4* allele to those without the allele. We calculated the population attributable fractions and confidence intervals using the AF package in R using the case control option (40, 41).

## Results

### Categorization of summary cognition status

Analyses were performed in both European (n=8399) and African (n=1605) ancestry groups. Participants with an “unclassified” summary cognition status were excluded from the analysis sample. Participants with missing *APOE* information were excluded from the study sample (**Supplementary Figure 1)**. In our total analytic sample (n=10004), 995 participants were classified with dementia (n_European_=724, n_African_=271) and 1061 participants were classified with CIND (n_European_=711, n_African_=350) (**Supplementary Table 3**). We also classified 1568 participants with borderline CIND (n_European_=1256, n_African_=312) and 6380 participants with normal cognition (n_European_=5708, n_African_=672). The proportion of cases within each cognition status was different by ancestry (*P <* 0.001).

### Distribution of polygenic scores (AD PGS)

AD PGSs were normally distributed within ancestries. We observed that PGS for AD, with and without the *APOE* gene region, differed by summary cognition status in both ancestries (all *P <* 0.007) with the lowest mean AD PGS in those with normal cognition in both ancestries. (**Supplementary Figure 2**).

### Univariate and bivariate analysis

The majority of the participants in each ancestry group were women n_European_=4786 (57.0%); n_African_=1013 (63.1%). The average age at the individual’s last cognition visit was 75.3 years (SD =9.04) in the European ancestry group and 72.2 years (SD=8.83) in the African ancestry group. There were 2800 participants with at least one copy of the *ε4* allele, n_European_=2208 (26.3%); n_African_=592 (36.9%). The year of last cognitive visit was 2014 for the majority of the analytic sample (n_European_=6285 (74.8%); n_African_=1265 (78.8%)). Sex, age, education, year of last visit, and *APOE-ε4* status differed by ancestry (*P <* 0.01). Hypertension, diabetes, smoking, alcohol use, and stroke status also differed by ancestry (*p*≤ 0.001). Depression status did not differ by ancestry (*p*=0.99).

Within the European ancestry sample, there were differences in demographic characteristics by summary cognition status (dementia, CIND, borderline CIND, normal) (**Table 1**). The mean age at last visit of those with dementia (84.2 years; SD=7.73) was higher than those with normal cognition (72.9 years; SD=8.17) (*P <* 0.001). The mean years of education were lower in those with dementia (11.9 years; SD=2.90), compared to those with normal cognition (13.7 years; SD=2.31). The distribution of year of last visit differed by cognition status (*P <* 0.001). BMI, history of hypertension, diabetes, stroke, depression, smoking status, and alcohol use differed significantly across cognition category (*P <* 0.001) with higher proportions of hypertension, diabetes, stroke, and depression status in those with impaired cognition compared to normal cognition. Lower mean BMI, less alcohol use, and lower proportions of current smokers were observed in those with impaired cognition compared to normal cognition. *APOE-ε4* status differed by cognitive status in the European ancestry group (*P <* 0.001) only.

**Table 1.** Bivariate analyses of covariates by cognition status stratified by race among Health and Retirement study (HRS) participants with core measurements taken from 1995-2014. The analytic sample includes participants at least three visits of cognition measured from ages 60 and older with an assigned cognition status, and complete genetic information. Analysis was split by genetic ancestry determined by principal component analysis: European ancestry (n=8399) and African ancestry (n=1605). Associations by race between cognition status (normal, borderline cognitively impaired-no dementia (CIND), CIND, and dementia) were tested by ancestry. Categorical variables are represented by n (%) with Chi-square test for association. Continuous variables are represented by mean (SD) with ANOVA test for association.

|  | European Ancestry (n=8399) |  |  |  |  | African Ancestry (n=1605) |  |  |  |  |
| --- | --- | --- | --- | --- | --- | --- | --- | --- | --- | --- |
|  | Normal<br>n=5708 | Borderline<br>CIND<br>n=1256 | CIND<br>n=711 | Dementia<br>n=724 | P-value | Normal<br>n=672 | Borderline<br>CIND<br>n=312 | CIND<br>n=350 | Dementia<br>n=271 | P-value |
| AD polygenic score (no APOE)<sup>a</sup> | -0.03 (1.01) | 0.09 (0.98) | 0.02 (1.00) | 0.03 (0.99) | 0.003 | -0.10 (1.01) | 0.00 (1.03) | -0.02 (0.96) | 0.15 (0.96) | 0.007 |
| AD polygenic score (with APOE) | -0.03 (1.00) | 0.10 (0.99) | 0.03 (0.98) | 0.06 (1.01) | <0.001 | -0.10 (1.01) | 0.00 (1.03) | -0.02 (0.96) | 0.15 (0.96) | 0.005 |
| APOE variant status<sup>b</sup> |  |  |  |  | - |  |  |  |  | - |
| ε2/ε2 | 29 (0.51%) | 12 (0.96%) | 5 (0.70%) | 1 (0.14%) |  | 6 (0.89%) | 5 (1.60%) | 7 (2.00%) | 2 (0.74%) |  |
| ε2/ε3 | 780 (13.7%) | 144 (11.5%) | 83 (11.7%) | 72 (9.94%) |  | 89 (13.2%) | 41 (13.1%) | 49 (14.0%) | 40 (14.8%) |  |
| ε2/ε4 | 117 (2.05%) | 23 (1.83%) | 17 (2.39%) | 26 (3.59%) |  | 23 (3.42%) | 19 (6.09%) | 16 (4.57%) | 19 (7.01%) |  |
| ε3/ε3 | 3528 (61.8%) | 738 (58.8%) | 408 (57.4%) | 391 (54.0%) |  | 337 (50.1%) | 153 (49.0%) | 173 (49.4%) | 111 (41.0%) |  |
| ε3/ε4 | 1161 (20.3%) | 306 (24.4%) | 183 (25.7%) | 211 (29.1%) |  | 188 (28.0%) | 80 (25.6%) | 95 (27.1%) | 85 (31.4%) |  |
| ε4/ε4 | 93 (1.63%) | 33 (2.63%) | 15 (2.11%) | 23 (3.18%) |  | 29 (4.32%) | 14 (4.49%) | 10 (2.86%) | 14 (5.17%) |  |
| APOE-ε4 binary status<sup>c</sup> |  |  |  |  | <0.001 |  |  |  |  | 0.093 |
| No ε4 allele | 4337 (76.0%) | 894 (71.2%) | 496 (69.8%) | 464 (64.1%) |  | 432 (64.3%) | 199 (63.8%) | 229 (65.4%) | 153 (56.5%) |  |
| ε4 allele present | 1371 (24.0%) | 362 (28.8%) | 215 (30.2%) | 260 (35.9%) |  | 240 (35.7%) | 113 (36.2%) | 121 (34.6%) | 118 (43.5%) |  |
| Age at last visit | 72.9 (8.17) | 78.1 (8.54) | 80.8 (8.40) | 84.2 (7.73) | <0.001 | 68.4 (6.84) | 71.4 (7.76) | 75.1 (8.94) | 78.8 (9.16) | <0.001 |
| Year of last visit |  |  |  |  | <0.001 |  |  |  |  | <0.001 |
| 2006 | 122 (2.1%) | 40 (3.2%) | 36 (5.1%) | 17 (2.4%) |  | 8 (3.0%) | 10 (2.9%) | 6 (1.9%) | 9 (1.3%) |  |
| 2008 | 318 (5.6%) | 116 (9.2%) | 65 (9.1%) | 66 (9.1%) |  | 32 (11.8%) | 32 (9.1%) | 11 (3.5%) | 20 (3.0%) |  |
| 2010 | 274 (4.8%) | 108 (8.6%) | 68 (9.6%) | 103 (14.2%) |  | 29 (10.7%) | 34 (9.7%) | 11 (3.5%) | 27 (4.0%) |  |
| 2012 | 347 (6.1%) | 165 (13.1%) | 105 (14.8%) | 164 (22.7%) |  | 39 (14.4%) | 25 (7.1%) | 19 (6.1%) | 28 (4.2%) |  |
| 2014 | 4647 (81.4%) | 827 (65.8%) | 437 (61.5%) | 374 (51.7%) |  | 163 (60.2%) | 249 (71.1%) | 265 (84.9%) | 588 (87.5%) |  |
| Sex |  |  |  |  | 0.004 |  |  |  |  | 0.067 |
| Male | 2403 (42.1%) | 582 (46.3%) | 333 (46.8%) | 295 (40.7%) |  | 226 (33.6%) | 116 (37.2%) | 147 (42.0%) | 103 (38.0%) |  |
| Female | 3305 (57.9%) | 674 (53.7%) | 378 (53.2%) | 429 (59.3%) |  | 446 (66.4%) | 196 (62.8%) | 203 (58.0%) | 168 (62.0%) |  |
| Education years | 13.7 (2.31) | 12.6 (2.37) | 11.6 (2.70) | 11.9 (2.90) | <0.001 | 13.3 (2.35) | 12.2 (2.37) | 10.3 (2.90) | 9.49 (3.82) | <0.001 |
| Cohort: |  |  |  |  | <0.001 |  |  |  |  | <0.001 |
| AHEAD<sup>e</sup> | 371 (6.50%) | 212 (16.9%) | 176 (24.8%) | 275 (38.0%) |  | 12 (1.79%) | 11 (3.53%) | 38 (10.9%) | 54 (19.9%) |  |
| CODA<sup>f</sup> | 428 (7.50%) | 172 (13.7%) | 126 (17.7%) | 147 (20.3%) |  | 5 (0.74%) | 10 (3.21%) | 34 (9.71%) | 30 (11.1%) |  |
| Remaining HRS cohorts | 4909 (86.0%) | 872 (69.4%) | 409 (57.5%) | 302 (41.7%) |  | 655 (97.5%) | 291 (93.3%) | 278 (79.4%) | 187 (69.0%) |  |
| BMI (kg/m<sup>2</sup>) at last visit | 29.0 (6.14) | 28.2 (6.11) | 27.3 (6.22) | 25.5 (5.36) | <0.001 | 31.5 (7.31) | 30.7 (7.13) | 30.2 (7.48) | 27.7 (6.49) | <0.001 |
| Ever hypertension |  |  |  |  | <0.001 |  |  |  |  | <0.001 |
| No | 2090 (36.6%) | 330 (26.3%) | 186 (26.2%) | 190 (26.2%) |  | 134 (19.9%) | 61 (19.6%) | 43 (12.3%) | 29 (10.7%) |  |
| Yes | 3618 (63.4%) | 926 (73.7%) | 525 (73.8%) | 534 (73.8%) |  | 538 (80.1%) | 251 (80.4%) | 307 (87.7%) | 242 (89.3%) |  |
| Diabetes status |  |  |  |  | <0.001 |  |  |  |  | 0.005 |
| No | 4420 (77.4%) | 910 (72.5%) | 488 (68.6%) | 540 (74.6%) |  | 437 (65.0%) | 180 (57.7%) | 198 (56.6%) | 148 (54.6%) |  |
| Yes | 1288 (22.6%) | 346 (27.5%) | 223 (31.4%) | 184 (25.4%) |  | 235 (35.0%) | 132 (42.3%) | 152 (43.4%) | 123 (45.4%) |  |
| Stroke status |  |  |  |  | <0.001 |  |  |  |  | <0.001 |
| No | 5303 (92.9%) | 1074 (85.5%) | 576 (81.0%) | 508 (70.2%) |  | 620 (92.3%) | 282 (90.4%) | 286 (81.7%) | 184 (67.9%) |  |
| Yes | 405 (7.10%) | 182 (14.5%) | 135 (19.0%) | 216 (29.8%) |  | 52 (7.74%) | 30 (9.62%) | 64 (18.3%) | 87 (32.1%) |  |
| Depression Status |  |  |  |  | <0.001 |  |  |  |  | <0.001 |
| Low CESD<sup>d</sup> | 3996 (70.0%) | 700 (55.7%) | 360 (50.6%) | 361 (49.9%) |  | 477 (71.0%) | 206 (66.0%) | 169 (48.3%) | 123 (45.4%) |  |
| High CESD | 1712 (30.0%) | 556 (44.3%) | 351 (49.4%) | 363 (50.1%) |  | 195 (29.0%) | 106 (34.0%) | 181 (51.7%) | 148 (54.6%) |  |
| Smoking status |  |  |  |  | <0.001 |  |  |  |  | 0.434 |
| Never | 2509 (44.0%) | 503 (40.0%) | 267 (37.6%) | 329 (45.4%) |  | 288 (42.9%) | 129 (41.3%) | 129 (36.9%) | 108 (39.9%) |  |
| Former | 2686 (47.1%) | 621 (49.4%) | 365 (51.3%) | 365 (50.4%) |  | 289 (43.0%) | 131 (42.0%) | 168 (48.0%) | 128 (47.2%) |  |
| Current | 513 (8.99%) | 132 (10.5%) | 79 (11.1%) | 30 (4.14%) |  | 95 (14.1%) | 52 (16.7%) | 53 (15.1%) | 35 (12.9%) |  |
| Alcohol status |  |  |  |  | <0.001 |  |  |  |  | <0.001 |
| No | 2420 (42.4%) | 725 (57.7%) | 488 (68.6%) | 565 (78.0%) |  | 395 (58.8%) | 198 (63.5%) | 254 (72.6%) | 220 (81.2%) |  |
| Yes | 3288 (57.6%) | 531 (42.3%) | 223 (31.4%) | 159 (22.0%) |  | 277 (41.2%) | 114 (36.5%) | 96 (27.4%) | 51 (18.8%) |  |

Acronyms: **CIND**: Cognitive impairment – no dementia; **AHEAD**: Asset and Health Dynamics among the Oldest Old; **CODA**: Children of the Depression study; **CESD**: Center for Epidemiologic Studies for Depression Scale; **SD**: standard deviation
a: Weights derived from Kunkle (IGAP, 2019) (25);
b: *APOE* status was genotyped using two SNPs (rs7412 and rs429358) resulting in three alleles of *APOE*. The frequencies listed are of the possible allelic combinations
c: Binary status was determined by the presence of the ε4 allele in participants
d: Binary cutoff determined by ancestry specific median of all CESD measures prior to last cognitive assessment

Likewise, in the African ancestry sample, the mean age at last visit of those with dementia (78.8; SD=9.16) was higher than those with normal cognition (68.4; SD=6.84) (*P <* 0.001). The mean years of education were lower among those with dementia (9.49 years; SD=3.82), relative to those with normal cognition (13.3 years; SD=2.35). The distribution of year of last visit differed by cognition status (*P <* 0.001).BMI, history of hypertension, diabetes, stroke, depression, and alcohol use differed significantly across cognition category (*P <* 0.01) with higher proportions of hypertension, diabetes, stroke, and depression status in those with impaired cognition compared to normal cognition. Higher mean BMI and more alcohol use were observed in those with normal cognition relative to impaired cognition.

### PGS association with dementia outcomes

Logistic regression models were run separately by ancestry **(Table 2**). In the European ancestry sample, age and education (Model 1) were associated with each impaired cognitive status relative to normal cognition (*P <* 0.0001). Sex was not associated with dementia relative to normal cognition (*p*=0.06) in Model 1, but was associated with CIND and borderline CIND relative to normal cognition (both *P <* 2 × 10^−5^). AD PGS, without the *APOE* gene region, was associated with dementia compared to normal cognition (Model 2a: OR=1.13, 95% CI: 1.03, 1.24). *APOE-ε4* status was also associated with dementia compared to normal cognition (Model 2b: OR=2.46, 95% CI: 2.02, 2.99). In Model 3, both AD PGS and *APOE-ε4* status were significantly and independently associated with dementia relative to normal cognition. After adjusting for age, sex, education, and *APOE-ε4* status, a one standard deviation increase in AD PGS was associated with a 1.10 (95% CI: 1.00, 1.20) times higher odds of dementia relative to normal cognition in European ancestry. After adjusting for age, sex, education, year at last visit, and AD PGS, carrying an *APOE-ε4* allele was associated with 2.42 (95% CI: 1.99, 2.95) times higher odds of dementia, relative to normal cognition in European ancestry. The association between *APOE-ε4* and the odds of dementia relative to normal cognition remained robust with the addition of health behaviors into the model (Model 4; OR 2.30 95% CI: 1.86, 2.85). The effect of AD PGS was consistent in magnitude, but became non-significant (OR 1.10 95% CI: 0.99, 1.21) with health behavior adjustment.

**Table 2.** Odds ratios (OR) of cognitive status, relative to normal status, explained by one standard deviation increase of polygenic score and the presence of an *APOE-ε4* allele in participants in the Health and Retirement study (HRS), of European and African ancestries, adjusted for age, sex, education, year of last visit, genetic principal component 1, and genetic principal component 2. Final model was additionally adjusted for by BMI, hypertension, diabetes, stroke, depression, smoking, and alcohol. Logistic regressions were performed on data subset.

|  | N | OR | Model 2a |  | OR | Model 2b |  | OR | Model 3 |  | OR | Model 4 |  |
| --- | --- | --- | --- | --- | --- | --- | --- | --- | --- | --- | --- | --- | --- |
|  |  |  | 95% CI | P-value |  | 95% CI | P-value |  | 95% CI | P-value |  | 95% CI | P-value |
| **European ancestry** |  |  |  |  |  |  |  |  |  |  |  |  |  |
| Polygenic score |  |  |  |  |  |  |  |  |  |  |  |  |  |
| Normal | 5708 | ref |  |  |  |  |  | ref |  |  | ref |  |  |
| Borderline CIND | 1256 | 1.15 | (1.08, 1.23) | <.0001 | - | - | - | 1.14 | (1.07, 1.21) | <.0001 | 1.13 | (1.06, 1.21) | <.0001 |
| CIND | 711 | 1.06 | (0.97, 1.16) | 0.188 | - | - | - | 1.05 | (0.96, 1.14) | 0.298 | 1.05 | (0.96, 1.15) | 0.288 |
| Dementia | 724 | 1.13 | (1.03, 1.24) | 0.008 | - | - | - | 1.10 | (1.00, 1.20) | 0.049 | 1.10 | (0.99, 1.21) | 0.07 |
| *APOE-ε4* |  |  |  |  |  |  |  |  |  |  |  |  |  |
| Normal | 5708 |  |  |  | ref |  |  | ref | - | - | Ref | - | - |
| Borderline CIND | 1256 | - | - | - | 1.46 | (1.26, 1.69) | <.0001 | 1.43 | (1.24, 1.65) | <.0001 | 1.45 | (1.25, 1.68) | <.0001 |
| CIND | 711 | - | - | - | 1.74 | (1.43, 2.11) | <.0001 | 1.73 | (1.43, 2.10) | <.0001 | 1.70 | (1.39, 2.07) | <.0001 |
| Dementia | 724 | - | - | - | 2.46 | (2.02, 2.99) | <.0001 | 2.42 | (1.99, 2.95) | <.0001 | 2.30 | (1.86, 2.85) | <.0001 |
| **African ancestry** |  |  |  |  |  |  |  |  |  |  |  |  |  |
| Polygenic score |  |  |  |  |  |  |  |  |  |  |  |  |  |
| Normal | 672 | ref |  |  | - | - | - | ref |  |  | ref |  |  |
| Borderline CIND | 312 | 1.06 | (0.87, 1.29) | 0.562 | - | - | - | 1.06 | (0.87, 1.30) | 0.555 | 1.07 | (0.86, 1.28) | 0.519 |
| CIND | 350 | 0.96 | (0.76, 1.20) | 0.702 | - | - | - | 0.96 | (0.76, 1.20) | 0.706 | 0.92 | (0.74, 1.16) | 0.46 |
| Dementia | 271 | 1.29 | (0.98, 1.70) | 0.072 | - | - | - | 1.29 | (0.97, 1.70) | 0.076 | 1.25 | (0.95, 1.65) | 0.141 |
| *APOE-ε4* |  |  |  |  |  |  |  |  |  |  |  |  |  |
| Normal | 672 | - | - | - | ref |  |  | ref |  |  | ref |  |  |
| Borderline CIND | 312 | - | - | - | 1.10 | (0.82, 1.47) | 0.541 | 1.10 | (0.82, 1.47) | 0.534 | 1.12 | (0.84, 1.51) | 0.443 |
| CIND | 350 | - | - | - | 1.08 | (0.78, 1.50) | 0.644 | 1.08 | (0.78, 1.50) | 0.648 | 1.14 | (0.81, 1.61) | 0.45 |
| Dementia | 271 |  |  |  | 1.77 | (1.20, 2.61) | 0.004 | 1.77 | (1.20, 2.61) | 0.004 | 1.74 | (1.15, 2.63) | 0.009 |

Model 2a: $\beta_0 + \beta_1(\text{Polygenic score}) + \beta_2(\text{age at last visit}) + \beta_3(\text{sex}) + \beta_4(\text{educational attainment}) + \beta_5(\text{year of last visit}) + \beta_6(\text{PC1}) + \beta_7(\text{PC2})$
Model 2b: $\beta_0 + \beta_1(\text{APOE-}\epsilon 4) + \beta_2(\text{age at last visit}) + \beta_3(\text{sex}) + \beta_4(\text{educational attainment}) + \beta_5(\text{year of last visit}) + \beta_6(\text{PC1}) + \beta_7(\text{PC2})$
Model 3: $\beta_0 + \beta_1(\text{Polygenic score}) + \beta_2(\text{APOE-}\epsilon 4) + \beta_3(\text{age at last visit}) + \beta_4(\text{sex}) + \beta_5(\text{educational attainment}) + \beta_6(\text{year of last visit}) + \beta_7(\text{PC1}) + \beta_8(\text{PC2})$
Model 4: $\beta_0 + \beta_1(\text{Polygenic score}) + \beta_2(\text{APOE-}\epsilon 4) + \beta_3(\text{age}) + \beta_4(\text{sex}) + \beta_5(\text{educational attainment}) + \beta_6(\text{year of last visit}) + \beta_7(\text{PC1}) + \beta_8(\text{PC2}) + \beta_9(\text{BMI}) + \beta_{10}(\text{hypertension}) + \beta_{11}(\text{diabetes}) + \beta_{12}(\text{stroke}) + \beta_{13}(\text{depression}) + \beta_{14}(\text{smoking}) + \beta_{15}(\text{alcohol})$
Acronyms: **HRS**: Health and Retirement Study; **BMI**: Body Mass Index; **OR**: odds ratio; **CI**: confidence interval; **CIND**: Cognitive impairment – no dementia

In the African ancestry sample, age and education (Model 1) were associated with each abnormal cognitive status relative to normal cognition (*P <* 0.003). Sex was not associated with any of the impaired cognitive statuses relative to normal cognition. AD PGS, without the *APOE* gene region, was not associated with dementia compared to normal cognition (Model 2a: OR=1.29, 95% CI: 0.98, 1.70). *APOE-ε4* status was associated with dementia compared to normal cognition (Model 2b: OR=1.77, 95% CI: 1.20, 2.61). In Model 3, *APOE-ε4* status remained significantly associated with dementia relative to normal cognition. After adjusting for age, sex, education, and AD PGS, carrying an *APOE-ε4* allele was associated with 2.10 (95% CI: 1.34, 3.28) times higher odds of dementia, relative to normal cognition.

### Sensitivity analyses

To compare the primary AD PGS without the *APOE* gene region to the AD PGS with the *APOE* gene region included, we tested for correlation between the two PGSs. Pearson correlation between these two AD PGSs (ρ_European_ = 0.9772, *P <* 0.0001; ρ_African_ = 0.9981, *P <* 0.0001). Thus, we did not include a separate set of analyses of an AD PGS with the *APOE* gene region.

To examine the effect of the P-value threshold selection for SNP inclusion in the PGS, we compared performance of PGSs developed using variable P-value thresholds (pT=1, 0.3, 0.1, 0.05, 0.01, 0.001) in Model 2a (logistic regression adjusting for age, sex, education, year at last visit, two genetic principal components, and AD PGS). Correlations between AD PGSs at different P-value threshold cutoffs ranged from 0.30 to 0.98 in the European ancestry sample and from 0.41 to 0.99 in the African ancestry sample (**Supplementary Table 4**). In the European ancestry sample, for the borderline CIND and CIND vs normal cognition models, we found no substantive difference in the association between AD PGSs at different thresholds and the outcome (**Supplementary Table 5**). AD PGSs were all significantly associated with the odds of borderline CIND (*P <* 0.05), while AD PGS was not associated with the odds of CIND, both relative to normal cognition. For the model estimating the odds of dementia vs normal cognition, the AD PGS was significantly associated with the outcome at pT=0.01 and pT=0.001 (*P <* 0.01), but not associated at pTs>0.01. The African ancestry sample results with other P-value thresholds were consistent with the results presented at pT=0.01. That is, AD PGS was not associated with odds of impaired cognition, relative to normal cognition at any pT.

To account for potential survival bias in our study sample, the oldest HRS cohorts (AHEAD and CODA) were removed, dropping the sample size from n=10004 to n=7913 (n_European_ = 6492; n_African_= 1421). In this subset of the analytic sample for European ancestry, with both AD PGS and the *APOE-ε4* allele included in the model, we observed that a one standard deviation increase in AD PGS was not associated with the odds of dementia OR=1.05 (95% CI: 0.92, 1.20), relative to normal cognition **(Supplementary Table 6)**. The presence of any *APOE- ε4* alleles remained significantly associated with the odds of dementia relative to normal cognition OR=2.60 (95% CI: 2.00, 3.38). When AHEAD and CODA were removed from the African ancestry sample, the effect of any *APOE-ε4* allele attenuated somewhat (from 1.77 to 1.55), but remained significantly associated with the odds of dementia relative to normal cognition.

We tested for an interaction between having any copies of the *APOE-ε4* allele and the AD PGS (both including and excluding the *APOE* region). There was not an interaction between *APOE-ε4* and the PGS including the APOE region (OR_interaction_=1.09, *P*=0.34). There also was not an interaction between *APOE-ε4* and the PGS excluding *APOE* region (OR_interaction_=1.10, *P*=0.30). This suggests that the effect of *APOE-ε4* is only additive and not multiplicative to the PGS. This also suggests that the effect of the *APOE-ε4* allele is the same as the *APOE* region.

We assessed alternative categories of *APOE* status, including a three-level variable for number of copies of the *ε4* allele (0, 1, or 2) (**Supplementary Table 7)**. In analyses in the European ancestry, having one copy and having two copies of APOE-*ε4* compared to no copies both significantly increased the odds of impaired cognition over normal cognition. There was a significant increase in the odds of dementia with one and with two copies of the APOE-*ε4* allele compared to normal cognition and no copies of the APOE-*ε4* allele in the African ancestry sample. While this may indicate utility in modeling APOE-*ε4* as two indicators for one or two copies of an *ε4* allele, the relative prevalence of two copies of *ε4* limits the power of this analysis (<5% in each ancestry). However, the results were overall consistent with the findings when modeling any copies of *APOE-ε4* vs no copies.

To assess the robustness of our findings, we adjusted the models for additional dementia risk factors (BMI, hypertension, depression, diabetes, smoking, and stroke) (**Table 2; Model 4**). After accounting for health covariates, in both the European ancestry and African ancestry groups, AD PGS was non-significant in modelling the odds of dementia status relative to normal (European ancestry: OR=1.10, 95% CI: 0.99, 1.21; African ancestry: OR=1.25, 95% CI: 0.95, 1.65) (**Table 2**). The effect of *APOE*-*ε4* remained significantly associated with the odds of impaired cognition in all models in the European ancestry (*P <* 0.0001). After accounting for the health covariates, having any copies of the *APOE*-*ε4* remained significantly associated with the odds of dementia in the African ancestry sample (OR=1.74, 95% CI: 1.15, 2.63).

### Receiver Operating Characteristic Curve

To address the differences in dementia prediction ability and potential clinical relevance, we assessed AUC using ROC curves. Our base model in the European ancestry, Model 1, produced a c statistic of 0.87. Using Model 1 as a reference, adding AD PGS did not significantly improve model discrimination relative to Model 1 (c_difference_=0.001, 95% CI: −0.0006, 0.0018, *P*=0.30). However, adding *APOE-ε4* status did significantly increase the classification accuracy of the model (c_difference_=0.0075, 95% CI: 0.0037, 0.0114, *P*=.0001). As expected from these results, adding the AD PGS did not significantly improve the classification accuracy over the model already including *APOE-ε4* (Model 2b: c_Model2b_=0.87; c_difference_=0.0001, 95% CI: - 0.0006, 0.0008, *p*=0.77). In the African ancestry group, no models performed more accurately than Model 1 **(Supplementary Table 8; Figure 1)**.

**Figure 1.**
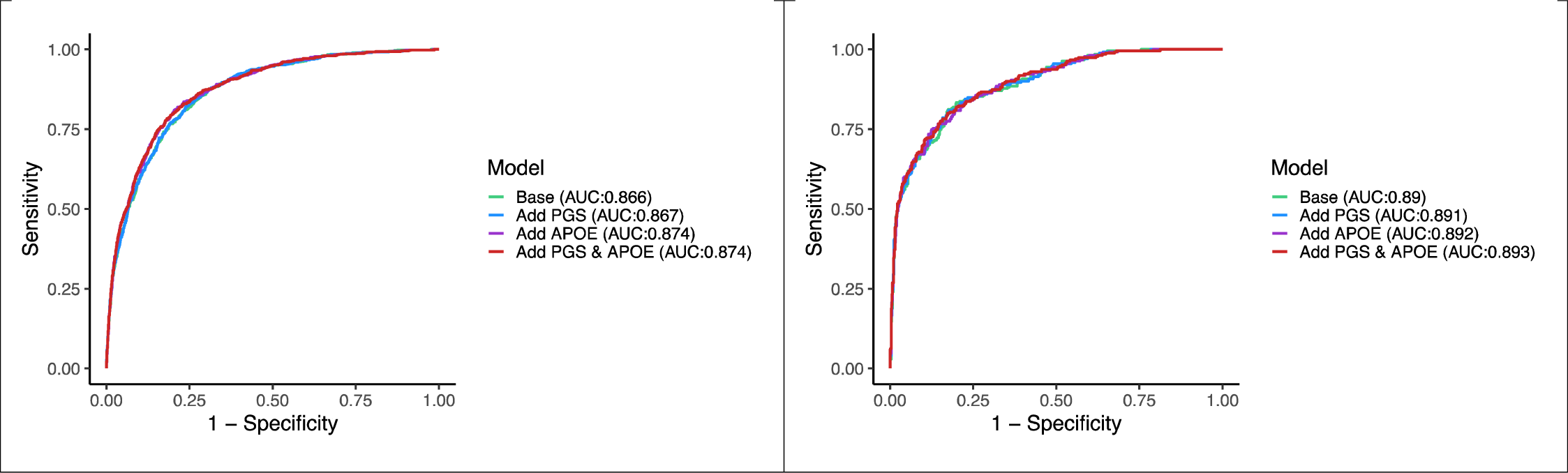
Receiver operating characteristic (ROC) curves for logistic regression models, looking at the association between polygenic risk score (PGS) and presence of *APOE-ε4* allele (APOE), with summary cognition statuses (dementia only) relative to normal status. This is among participants in the Health and Retirement Study (HRS) with more than two waves of cognition measured, no stroke, by ancestry (n_European_=8399; n_African_=1605)

### Attributable fraction

In models of European ancestry containing age at last cognition visit, sex, education, year of last cognition visit, two genetic principal components, *APOE-ε4* status, and the AD PGS (top 20% vs bottom 20%), 21.39% (95%CI 11.9%, 30.8%; *P*<.0001) of dementia cases and 9.58% (0.9%, 18.2%; *P*=.03) of borderline CIND cases can be attributed to having at least one copy of the *APOE-ε4* allele. Of borderline CIND cases, 18.99% (8.5%, 29.5%; *P*<.0001) can be attributed to being in the top 20% of the AD PGS distribution. In the African ancestry sample, 53.88% (35.7%, 72.0%; *P*=<.0001) of dementia cases can be attributed to being in the top 20% of the AD PGS distribution. All other attributable fractions were not significantly different than zero (**Supplementary Table 9**).

## Discussion

In this large US population-based study, we observed AD PGS and *APOE-ε4* status had independent effects on dementia compared to normal cognition in European ancestry after controlling for age and year at last visit, sex, educational attainment, and genetic principal components. The effect of the AD PGS on odds of dementia was suggestive, but the attenuation in effect size resulted in non-significant signal at this sample size after the addition of health-related AD risk factors. However the effect of any copies of an *APOE-ε4* allele remained robustly associated with the odds of dementia, relative to normal cognition. In the African ancestry sample, being a carrier for any copies of the *APOE-ε4* allele significantly increased the odds of dementia relative to normal cognition. In the African ancestry sample, we observed a higher magnitude of effect of the AD PGS than in the European ancestry sample, even while using weights from a European-based study of Alzheimer’s disease to build the PGS; however, with the smaller African ancestry sample size, we were less powered to detect a signal. Our study replicates previous *APOE* results and expands to also consider cumulative genetic risk, providing greater understanding of the genetic etiology of dementia.

### APOE and dementia

*APOE-ε4* is a consistent genetic risk factor associated with AD and related dementias but accounts for only a portion of the heritability of late-onset Alzheimer’s disease (42, 43). The dosage of *APOE-ε4* alleles may influence dementia risk; that is, having one copy of *ε4* may confer less risk than two copies of *ε4*. Having two copies of the *APOE-ε4* allele is rare, and many studies have examined at least one copy of *APOE-ε4* as a binary indicator due to sample size constraints. In supplemental analysis in this study, we detected dose-increasing odds by additional copy of *APOE-ε4* for dementia relative to normal cognition adjusting for base covariates and AD PGS, (one copy (n=237): OR=2.29, 95% CI: 1.87, 2.81; two copies (n=23): OR=4.93, 95% CI: 2.82, 8.62) in the European ancestry sample. We saw a similar pattern of the effect of one and two copies of *APOE-ε4* on the odds of dementia in the African Ancestry sample, though the observed effect size was somewhat smaller and we were again limited in our sample size (one copy (n=104): OR=1.68, 95% CI: 1.12, 2.51; two copies (n=14): OR=2.65, 95% CI: 1.11, 6.33).

### Polygenic risk and dementia

Several studies have examined the effect of AD PGS and AD or cognitive status, but these studies vary in their PGS creation techniques and modeling decisions. The majority of the studies have examined the AD PGS association on dementia or cognition were restricted to those of European ancestry or identifying as non-Hispanic Whites (43-48). A non-*APOE* PGS study (using a PGS from 19 SNPs outside the *APOE* gene) found that their PGS can successfully stratify *APOE-ε4* carriers into risk subgroups where the highest scores have a 62% increase in risk of late-onset Alzheimer’s disease over the lowest scores (49). Some non-APOE PGS have been used for AD-patient classification (50-54), with one reporting a 0.78 AUC (95%CI 0.77 to 0.80) (44) including age, sex, *APOE-ε4, APOE-ε2*, and the PGS for SNPs with AD association *p*-values <0.5. However, this report was using IGAP data from 2013 which included the study in which the AUC was calculated (GERAD). Our empirical estimate of the AUC for dementia in a European ancestry sample independent of the GWAS weights with similar covariates (any *APOE-ε4, APOE-ε2/ε2*, and the PGS for SNPs with AD association *p*-values <0.1) was higher at 0.85 (95%CI 0.83 to 0.86). This difference is likely due to the broader definition of our dementia phenotype. Other PGS analyses have reported AD-subtype discrimination from PGS created for AD (55), revealing multiple biological mechanisms underlying different AD subtypes. Importantly, we report similar analyses in an African ancestry sample though we strongly suggest that these analyses not be directly compared due to underlying genetic differences between ancestries.

A previous longitudinal analysis in the HRS (n=8253) featuring a 21 SNP AD PGS excluding *APOE* observed that a 0.1 unit increase in PGS was associated with 0.016 decreased memory score units (95% CI: −0.036 to 0.005) in European ancestry (n=7172) and 0.049 decreased memory score units (95% CI: −0.12 to 0.023) in African ancestry (N=1081) (56). In our study, we observed the effect of AD PGS on dementia risk in the same direction, providing evidence of the association of AD PGS on dementia or memory. We further extended our analysis, by including *APOE* genotype in the model as well adjusting for additional dementia risk factors. We also assessed the utility of transferring estimates from one population to another population. Allele frequencies, linkage disequilibrium patterns and the genetic architecture can vary by ancestral populations (27) based on recombination and demographic histories (57).

Participation or inclusion in the discovery GWAS is influenced by social and behavioral factors, which relates to the applicability of the discovery GWAS results to another population (58). Also, the background of non-genetic risk factors may be different across populations, likely affecting the observed genetic signal (58). Therefore, noting these limitations, we did not directly compare the genetic associations across groups (European ancestry and African ancestry), but focused on the within-ancestry findings and are cautious not to overstate our results. While trans-ancestry genetic analyses are challenging, it remains important to perform studies in multiple ancestries to demonstrate these fundamental differences, refine methods that generate AD PGSs, and call for more inclusive ancestry genome-wide studies.

Older participants with cognitive impairments may be more likely to die of other comorbid causes, and therefore capturing cognitively impaired cases in older age groups may be difficult (59). Therefore, individuals who have lived up to the age of 90 with no cognitive impairment, may skew our estimates of the odds of dementia. In addition, HRS samples for genetic analyses were collected starting in 2006 and *APOE* or other risk genotypes may have already begun to influence survival to that period of genetic data collection for inclusion in our analytic sample. In our primary analysis, we included the older age group (represented by the HRS cohorts, AHEAD and CODA). In sensitivity analyses, we removed these cohorts to observe if our results were robust to mortality selection. We lost over half (n=422, 58.3%) of our dementia cases removing AHEAD and CODA in the European ancestry sample. After removing the AHEAD and CODA cohorts, we observed one substantive change: the effect of PGS on the odds of dementia compared to normal cognition in the European ancestry sample (Model 3) attenuated from 1.10 (95%CI 1.00, 1.20, *P*=0.049) to 1.05 (95%CI 0.92, 1.20, *P*=0.447), while the OR for *APOE-ε4* in the same model increased in effect size and remained highly significant from 2.42 (95%CI 1.99, 2.95) to 2.60 (95%CI 2.00, 3.38) (*P*<.0001). Importantly, the odds ratios from each model were not significantly different from each other, indicating that mortality selection did not critically bias these results.

Unlike the process of clinical or pathological diagnosis of dementia and Alzheimer’s disease, dementia ascertainment in population cohorts can be highly variable. We used the Langa-Weir method that uses questions derived from the Telephone Interview of Cognitive Status, summed into a total score and assigned a cognition status using cut-points mirroring clinical diagnosis. Many dementia and cognition related studies have used the same approach (60, 61). Compared to other regression-based models, the Langa-Weir method has a comparable sensitivity and specificity (sensitivity: 75%; specificity: 83%) and provides balanced accuracy with prior clinical validation (62). Other known challenges such as time-varying biases and non-linear cognitive trajectory (63) were addressed by constructing a summary cognition status based on multiple visits. There are documented learning effects – where scores are higher the second time a participant sees a similar exam – for these cognition tests (64). Our summary cognition status excluded those with less than three visits, strengthening the cognitive status designation for each individual, and reducing practice effects.

### Clinical utility of polygenic risk scores

Assessing an ROC curve and calculating an AUC has allowed researchers to better understand how genetic scores can contribute to classification and potentially clinical diagnosis of complex disease outcomes. Many studies have seen some utility of using polygenic scores to help identify (for example) risk of cancer (65-68), multiple sclerosis (69), rheumatoid arthritis (70), Parkinson’s (71), and cardiovascular disease (72). AD PGSs have been investigated for their utility in stratifying individuals based on age of onset of AD, where individuals who had a top quartile AD PGS had an age of onset of 75 vs. an age of onset of 95 for the lowest quartile (73).

Despite the motivation to use AD PGS in clinical practice, scientists are understandably hesitant to encourage its use outside of research. The risk conferred by AD PGS is calculated at a population level and may not be appropriate to predict risk in an individual (74). In addition, current AD PGSs are not created with diverse populations as the reference weights, which may provide inaccurate risk for individuals of diverse ancestries (75), and clinical use in their present form may exacerbate health disparities (76). With improvement, ultimately, PGS may prove to be a useful tool in public health and preventative and therapeutic medicine. We may eventually be able to use PGSs for primary prevention (e.g. quantifying the genetic burden in subpopulations), secondary prevention (e.g. detecting high-risk individuals for disease screening), and tertiary prevention (e.g. a potential biomarker for optimizing treatment stratification) (77). However, PGSs as they are currently, were not designed to be used as a diagnostic tool nor are they sufficiently accurate enough for clinical diagnosis, particularly across ancestries.

## Data Availability

All data are publicly available https://hrs.isr.umich.edu/about

## Acknowledgements

We thank the participants in the Health and Retirement Study and the staff who catalogued their information. This work was supported by the National Institutes of Health (grant numbers R01 AG055406, R01 AG055654, R25 AG053227, R01 AG053972, R03 AG048806).

The Health and Retirement Study is sponsored by the National Institute on Aging (U01 AG009740) and is conducted at the Institute for Social Research, University of Michigan.

## Conflicts

The authors declare no conflicts of interest.

## Acronyms

AD: Alzheimer’s dementia
APOE: Apolipoprotein E
CI: Confidence interval
PGS: Polygenic risk score
GWAS: Genome wide significant studies
SNPs: Single nucleotide polymorphism
HRS: Health and retirement study
CIND: Cognitive impairment –
no dementia
PC: Principal component
IGAP: International genomics of Alzheimer’s project
BMI: Body mass index
SD: Standard deviation
df: degrees of freedom
AUC: Area under the curve

## Supplementary Material

**Supplementary Table 1**. Summary cognition status classification decision tree. Among participants in the Health and Retirement Study (HRS) with more than two wave specific dementia status values between the years 1995 – 2014 (n=10175), participants’ cumulative cognitive status were determined. The criteria column describes the specifications to inform the designation of the cognition status, found in the outcome decision column. Specific sample sizes for each cognition status (dementia, cognitively impaired no dementia (CIND), borderline CIND, normal cognition, and unclassified) are found in the subsequent columns.

**Supplementary Table 3**. Univariate analyses of covariates among Health and Retirement study (HRS) participants with core measurements taken from 1995-2014. The study sample consists of participants with genetic data collected during their time in HRS. The analytic sample further restricts to participants with at least three visits of cognition measured from ages 60 and above that were assigned a cognition status. Analysis was split by genetic ancestry determined by principal component analysis: European ancestry (n=8399) and African ancestry (n=1605). Categorical variables are represented by n (%) with Chi-square test for association. Continuous variables are represented by mean (SD) with ANOVA test for association.

**Supplemental Table 4**. Pearson’s correlation coefficients between Alzheimer’s disease polygenic scores (PGS) created from the Kunkle et al., 2019 Alzheimer’s GWAS meta-analysis Stage 1 at different P-value thresholds for SNPs included in the AD PGS (pT=1.0, 0.3, 0.1, 0.05, 0.01, 0.001). All these AD PGSs are created after removing the linkage disequilibrium block of the *APOE* gene (chr19:45384477-45432606, GRCh37/hg19) from the summary statistics. Upper triangle (gray) contains correlation coefficients from European ancestry, lower triangle (no shading) from African ancestry. All correlations are significantly different than 0 (*P <* .0001).

**Supplemental Table 5**. Odds ratios (OR) of cognitive status, relative to normal status, explained by one standard deviation increase of polygenic score (PGS) and the presence of a *APOE-ε4* allele in participants in the Health and Retirement study (HRS), of European and African ancestry, adjusted for age, sex, education, year of last cognition visit, and two genetic principal components. Examining different P-value thresholds for the Kunkle et al., 2019 GWAS meta-analysis Stage 1 for SNPs included in the AD PGS (pT=1.0, 0.3, 0.1, 0.05, 0.01, 0.001). Results shown are for Model 2a only.

**Supplemental Table 6**. Odds ratios (OR) of cognitive status, relative to normal status, explained by one standard deviation increase of polygenic score (PGS) and the presence of a *APOE-ε4* allele in participants in the Health and Retirement study (HRS), of European and African ancestry, adjusted for age, sex, education, year of last cognition visit, and two genetic principal components, excluding cases that are part of the AHEAD and CODA cohorts from the HRS. Logistic regressions were performed on data subsets by cognitive status, relative to normal cognition (ref).

**Supplemental Table 7**. Odds ratios (OR) of cognitive status, relative to normal status, explained by a one standard deviation increase of polygenic score (PGS) and *APOE-ε4* status based on *ε4* allele copies (0, 1, or 2 copies of *ε4*) of participants in the Health and Retirement study (HRS), of European and African ancestry, adjusted for age, sex, education, year of last cognition visit, and two genetic principal components. Final model was additionally adjusted for by BMI, diabetes, hypertension, smoking, and depression status. Logistic regressions were performed on data subsets based on cognition status, relative to normal cognition (ref).

**Supplementary Table 8**. Receiver operating characteristic (ROC) contrast statistics for logistic regression models, looking at the association between polygenic risk score (PGS) and presence of *APOE-ε4* allele (APOE), with summary cognition statuses (dementia only) relative to normal status. This is among participants in the Health and Retirement Study (HRS) with more than two waves of cognition measured at or after age 60, by ancestry (n_European_=6432 and n_African_=943).

**Supplementary Table 9**. Attributable fraction (AF) for logistic regression models, looking at the association between polygenic risk score (PGS) and presence of *APOE- ε4* allele (APOE), with summary cognition status relative to normal status. This is among participants in the Health and Retirement Study (HRS) with more than two waves of cognition measured at or after age 60, by ancestry: European and African ancestries. PGSs are dichotomized into top 20% and bottom 20% of Alzheimer’s disease polygenic score and *APOE-ε4* is modeled as any copy of *APOE-ε4* versus none. Crude models include only the variable of interest (AD PGS top 20% vs bottom 20% or *APOE-ε4* carrier) as predictors. Fully adjusted models include age at last cognition visit, sex, education, year of last cognition visit, two genetic principal components, *APOE-ε4* status, and the AD PGS (top 20% vs bottom 20%).

**Supplementary Figure 1.**
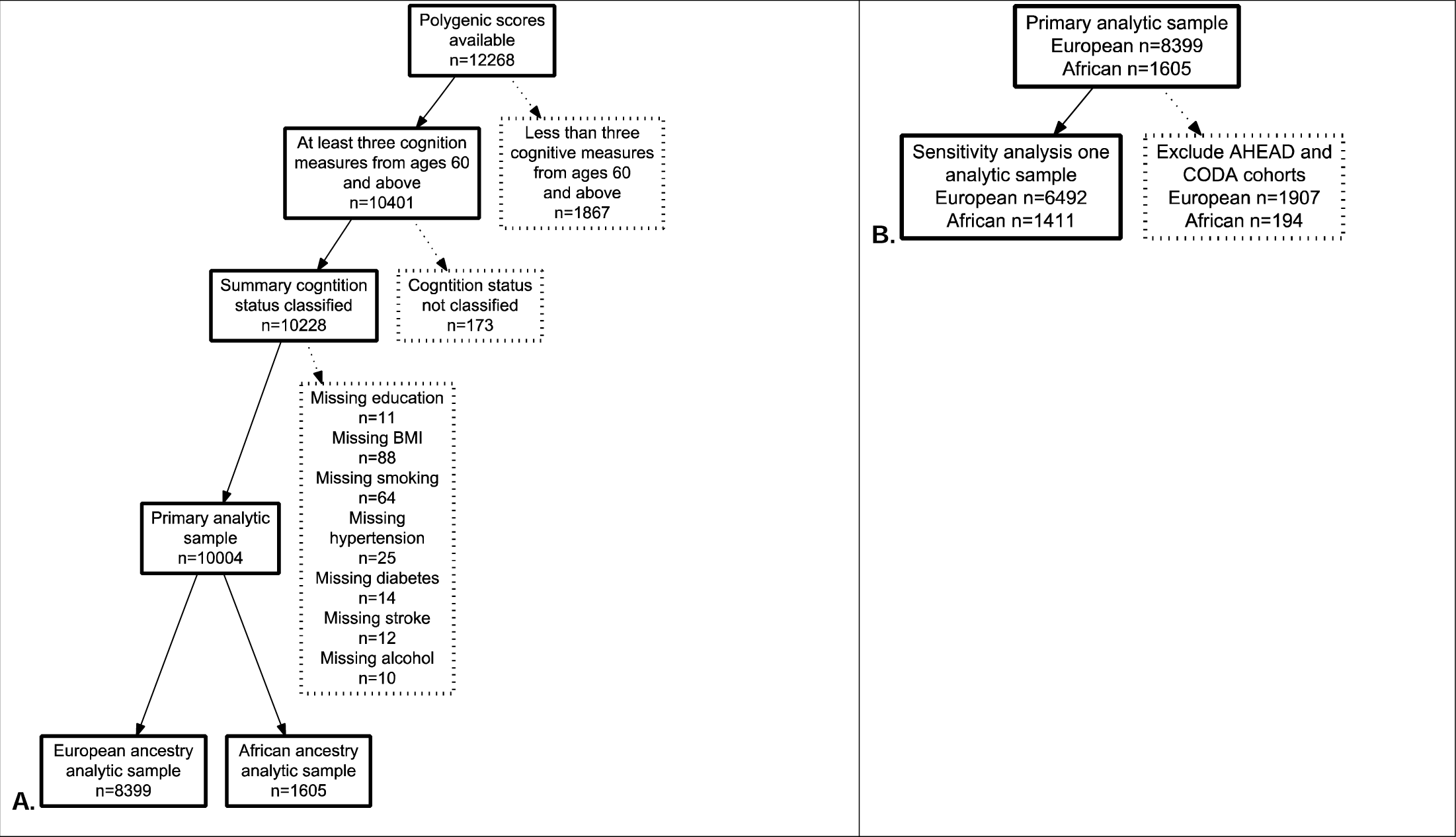
Analytic sample exclusion criteria diagram. The primary sample includes participants in the Health and Retirement Study (HRS) between the years 1995 – 2014 with complete genetic information. The primary analytic sample n=10004 included European ancestry n_European_=8399 and African ancestry n_African_=1605 cohorts (Panel A). A sensitivity analysis was performed by removing cases from the AHEAD and CODA cohorts (n_European_=6492 and n_African_=1411) (Panel B).

**Supplementary Figure 2.**
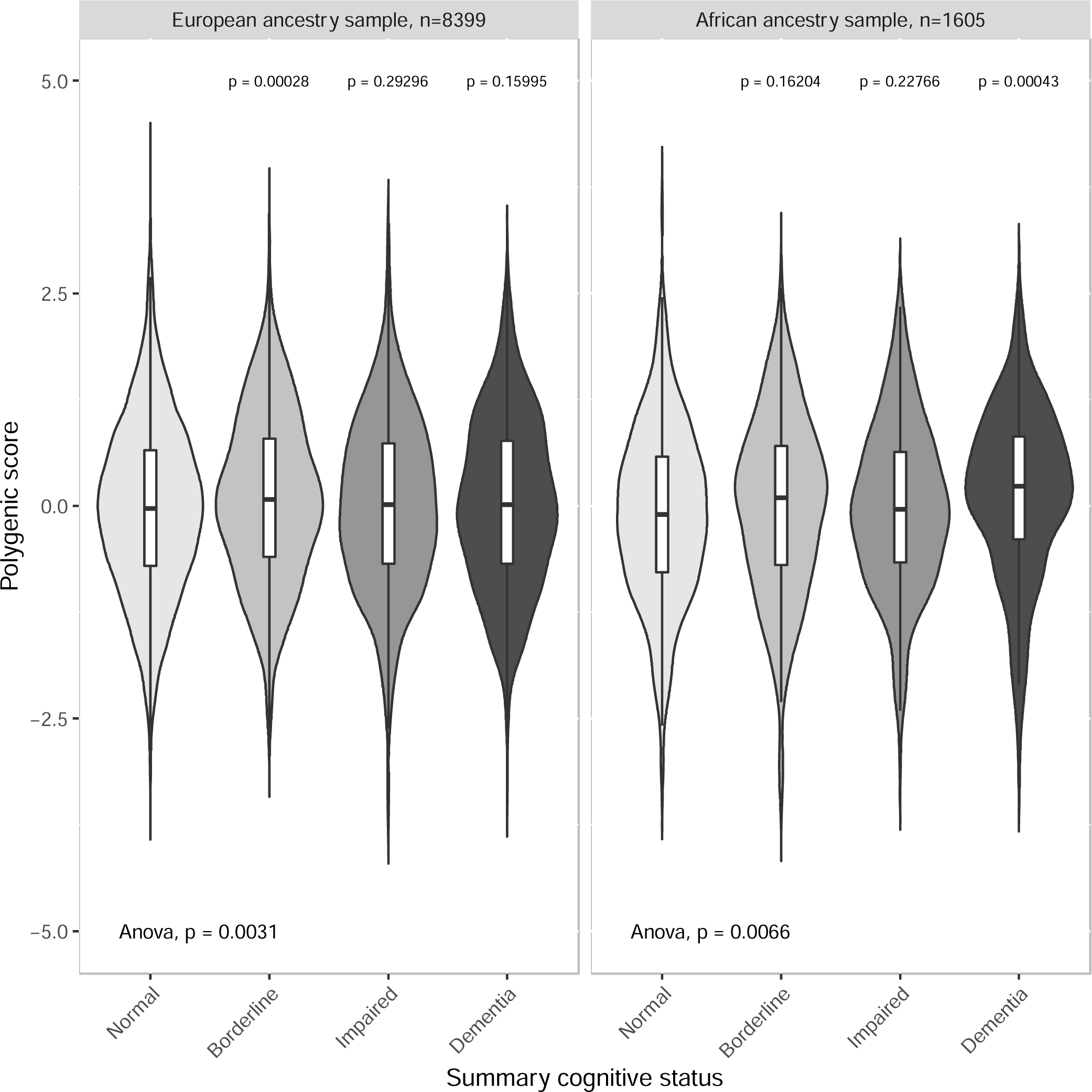
Polygenic score (PGS) distribution across summary cognition status (dementia, cognitive impairment-no dementia (CIND), borderline CIND, and normal), among participants in the Health and Retirement Study (HRS) with more than two waves of cognition measured, in both European and African ancestry participants. Pairwise t-tests were performed between cognition statuses, with dementia status as reference, *P-values* reported in plot. Global ANOVA tests were performed and reported in plot to test overall mean differences between each cognition status.

## Notes

### Competing Interest Statement

The authors have declared no competing interest.

